# Mobility resolution needed to inform predictive epidemic models for spatial transmission from mobile phone data

**DOI:** 10.1101/2024.10.11.24315335

**Authors:** Giulia Pullano, Shweta Bansal, Stefania Rubrichi, Vittoria Colizza

**Affiliations:** Department of Biology, Georgetown University, Washington, DC, USA; Orange Labs, Sociology and Economics of Network and Services (SENSE), Chatillon, France; Sorbonne Université, INSERM, Institut Pierre Louis d’Epidémiologie et de Santé Publique, IPLESP, Paris, France

## Abstract

Mobility flows extracted from mobile phone data have been extensively used in recent years to inform spatial epidemic models for the study of various infectious disease epidemics, including Malaria, Cholera, and Ebola. Most recently, the COVID-19 pandemic marked a historic shift, as it led to the sharing of unprecedented fine-scale mobility data. This abundancy of data illuminated the geographical variability in transmission patterns and underscored the importance of the use of mobility data for public health questions. Little attention has been devoted however to (i) the definition of the mobility process that is relevant to the epidemic spread, and (ii) the mobility data resolution that is needed to describe the invasion dynamics. We take advantage of a real-world dataset, gathered from mobile phone users in Senegal to define three epidemiological couplings between locations, based on different characterizations of the mobility process, and at varying resolution levels. They are based respectively on: (i) the total number of displacements between any two municipalities on two consecutive calls (Displacement-based *D*); (ii) the number of calls made by residents in each location (Location-based *L*); (iii) the most visited location of residents during daytime (Most visited location-based *C*). To assess the impact of the different coupling definitions on the epidemic diffusion, we use them to inform mobility in a spatial epidemic model. We found that preserving any displacement on the observed trajectories from mobile phone data does not capture the epidemiological link between different locations, for infections where daily mobility is important (e.g. airborne or direct contact diseases). Most importantly, we found that at the country scale, places in which individuals spend most of their time including workplaces, schools or particular point of interests like restaurants or theater and are the dominant driver of disease diffusion. In fact, tracking in detail individual activities beyond home and all visited locations during the day does not add epidemiological important information. Novel paradigms for the release of mobile phone data to researchers can therefore be envisioned that strengthen privacy and confidentiality, while at the same time providing enough details - specifically aggregated home-visited locations coupling - to inform predictive epidemic models.

## Introduction

Fine-grained data on human mobility patterns have become key elements for building spatially explicit epidemic models^1–3^. In the last decades, researchers have largely used mobility fluxes extracted from many data sources including census data, surveys, transport statistics, commuting data^4,5^.

Recent digital innovation has led to an explosive data growth. Everyday human interactions with digital services and technologies like mobile phones generate massive geolocated datasets (e.g. mobile phone data, GPS, online data). These sources allow gaining insight into human behaviour at large scale^4,6,7^. In 2019, there were already around 5 billion unique mobile phone subscribers globally, which means a penetration rate of 67% of the global population^8^. This has created an impressive collection of individual digital tracks all over the world, and it made mobile phone an extremely rich and informative source of mobility data.

In recent years, mobile phone data have been validated as a good proxy for commuting network ^9^ and it has been largely used to inform spatially explicit epidemics models^9–14^. Epidemiological studies have been done on disparate scenarios like malaria^13,15^, cholera^12,16^, schistosomiasis^17^, dengue^18^, HIV^19,20^, 2013-2016 Ebola outbreak^21,22^. During the COVID-19 pandemic, mobile phones have been used for the first time for tracking human mobility and to inform models in near real-time ^23–29^. Given the emergency, we witnessed an unprecedented massive data sharing, and this opened new challenges in terms of their integration into models and minimization of shared information to protect mobile phone users privacy.

Mobile phone data are integrated into spatially-explicit epidemic models through coupling forces between any pair of locations. The coupling forces model the probability of spatial transmission among connected locations due to human mobility. Given the individual daily trajectories extracted from mobile phones, the methodological challenge is to translate such high-resolution individual information to coupling forces linking geographical areas. This means to i) define which locations need to be coupled and ii) quantify the probability of coupling between the connected locations.

So far, coupling forces have been extracted and integrated into epidemic models without pay attention to the accuracy of the process of aggregation of the mobility flows relevant to disease spatial propagation. Aggregating properly mobile phone traces means to assess how many details of them are needed to describe epidemiological links between locations, and which details are instead negligible, or noise.

Considering the privacy issues on dealing with mobile phone data and the time needed to extract and pre-processed them before their sharing with researchers^30–34^, to know a priori the resolution needed to inform an epidemic model become crucial in the management of a crisis. We first reviewed several methods commonly used in literature to aggregate mobile phone traces at different temporal and spatial resolution. We selected three main aggregation methods at maximum, medium and lowest resolution (**Figure 1**), and we proceed by evaluating the outcome of those on the modelled epidemic spreading. The first coupling matrix counts the number of displacements between any two consecutive calls made by a user (*Displacement-based coupling matrix D*)^11^. Accounting for the full trajectory of each individual, we referred to this as the high-resolution coupling. The second coupling matrix connects the residence location of each user to all their visited locations, with a coupling force that is proportional to the number of calls made in each location (*Location-based coupling matrix L*)^12^. The definition of the mobility process associated to this epidemiological coupling is radically different from the first one. In this case, the full trajectory is lost in favour of a coupling between the home and the visited locations. We refer to *L* as the medium-resolution coupling. The third coupling matrix exclusively connects the residence location of each user to their most visited location and it is computed on the number of calls^13^ (Most visited location-based *C*). Comparing the aggregation procedures, this approach is at the lowest resolution, as it considers only one visited location for each user.

**Figure 1.**
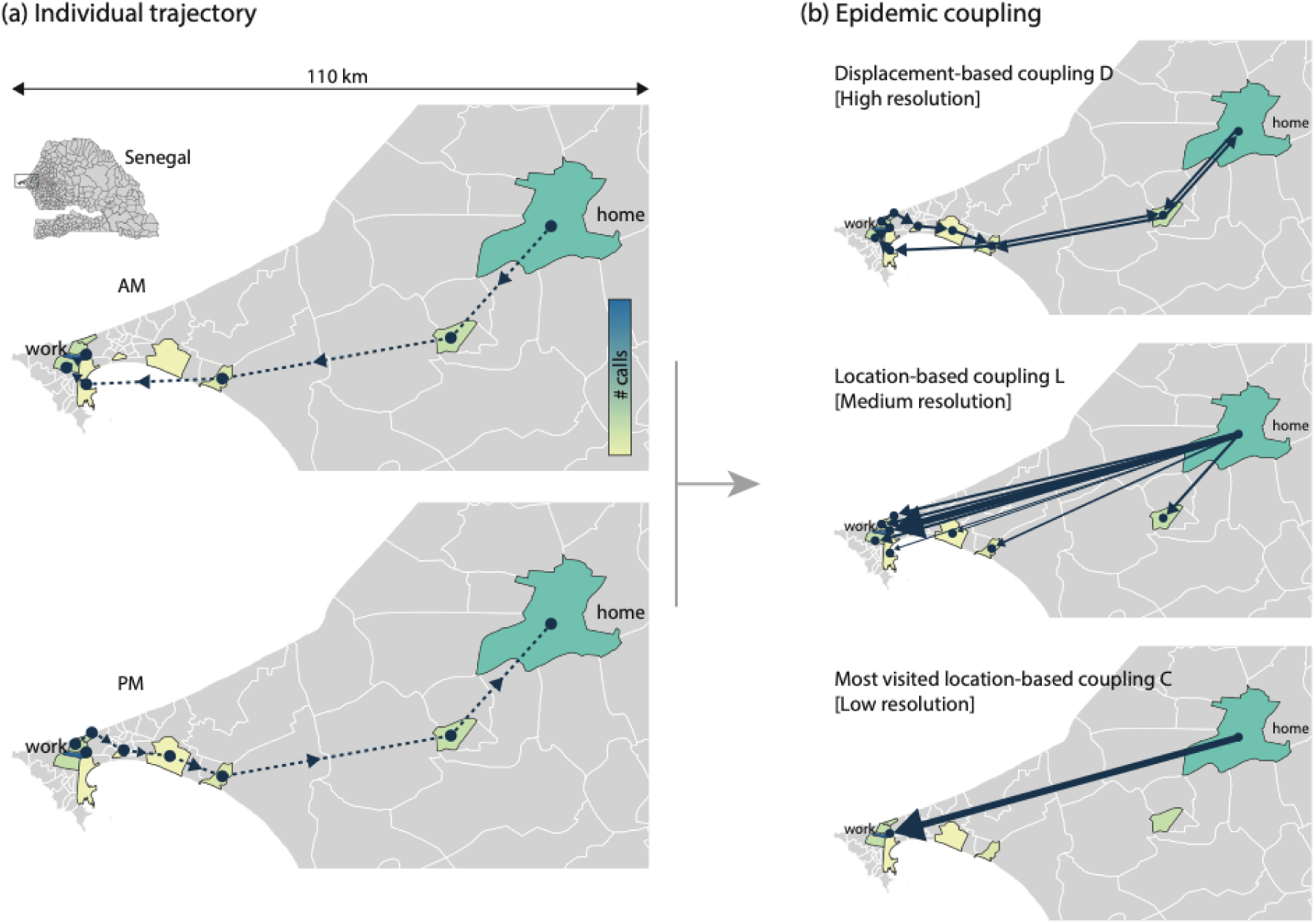
The aggregation processes. *D*) High-resolution method^11^; *L*) Medium-resolution method^12^; *C*) Lower-resolution method^44^.

Comparing *D* with *L*, we compared two different mobility definition. in *D* the probability of moving from one location to another and, in *L*, the probability of spending time in each location. Comparing *L* with *C*, instead, we explored the role of time spent in any place, accounting for the same transmissibility in each location.

We quantified the associated coupling matrices (*D, L, C*) for each month from January-December 2013 in Senegal (9,569,425 users). To assess the role of the mobility definition on disease diffusion, we integrated the estimated coupling matrices in a novel metapopulation model. We developed a metapopulation model that explicitly accounts for disease dispersal due to visitors to a location or to returning resident to their municipality of residence. The novelty in the model is the normalization factor of the force of infection that depends on the effective population in a given place, which is composed of all these three categories of people. We then used this model to simulate the spread of influenza-like and Ebola-like diseases and evaluated the outcomes of a stochastic SEIR dynamics. We thus evaluated how the simulated epidemic behaviour depends on the underlying spatial and time aggregation scheme, by investigating the time to the first infection in each location and the invasion epidemic paths from the seed.

## Results

### Senegalese spatial representation

The study was performed at municipality level, the 4th level of administrative division of the country. Basing on Census 2013 provided by the Senegalese national institute of statistics ANSD, we considered, 13092348 inhabitants distributed among 443 municipalities. We took into account only the municipalities covered by mobile phone antennas throughout 2013, and we selected 394 ones (80% of the whole municipalities in Senegal): 46 urban and 348 rural ones. The 46 urban municipalities are located in the following cities: Dakar, Guediawaye, Pikine, Rufisque, Thies.

### Statistical comparison of coupling matrices

Defining the mobility by three aggregation methods at different spatio-temporal resolutions (*D,L,C*), we extracted the three coupling matrices per month, and we obtained 12 directed networks for each method. Connections between links depend on the aggregation process, so the resulting networks have different topologies (**Figure 1**). To understand similarities and differences between any pair of networks, we measured their association under conditions of multicollinearity with a Multiple Regression Quadratic Assignment Procedure (MRQAP), and we found that all methods are highly correlated (regression coefficient *r_C_*_,*L*_ = 1, *r_D_*_,*L*_ = 0.99, *r_D_*_,*C*_ = 0.99). Even though all three matrices are correlated to each other, elements in *D* differ from the other two methods of an order of magnitude. While *C* and L have a quite similar probability distribution of the coupling probabilities (**Figure 3a**), in *D* the median of the distribution is around 1 order of magnitude lower (**Figure 2a**). Largest differences are on links that connect Urban/Rural municipalities (**Figure 2b**). Moreover, the median of the geographical distance distribution is lower in *D* compared with the others two methods (**Figure 2c**). **Figure 2b** shows also differences between *D, L* and *C* on the outgoing probability in any location. Such difference increases on municipalities far from the urban areas (**Figure 2d,e,f**). In this latter case, the outgoing probability in *D* is around two order of magnitude smaller compared with *L* and *C*.

**Figure 2.**
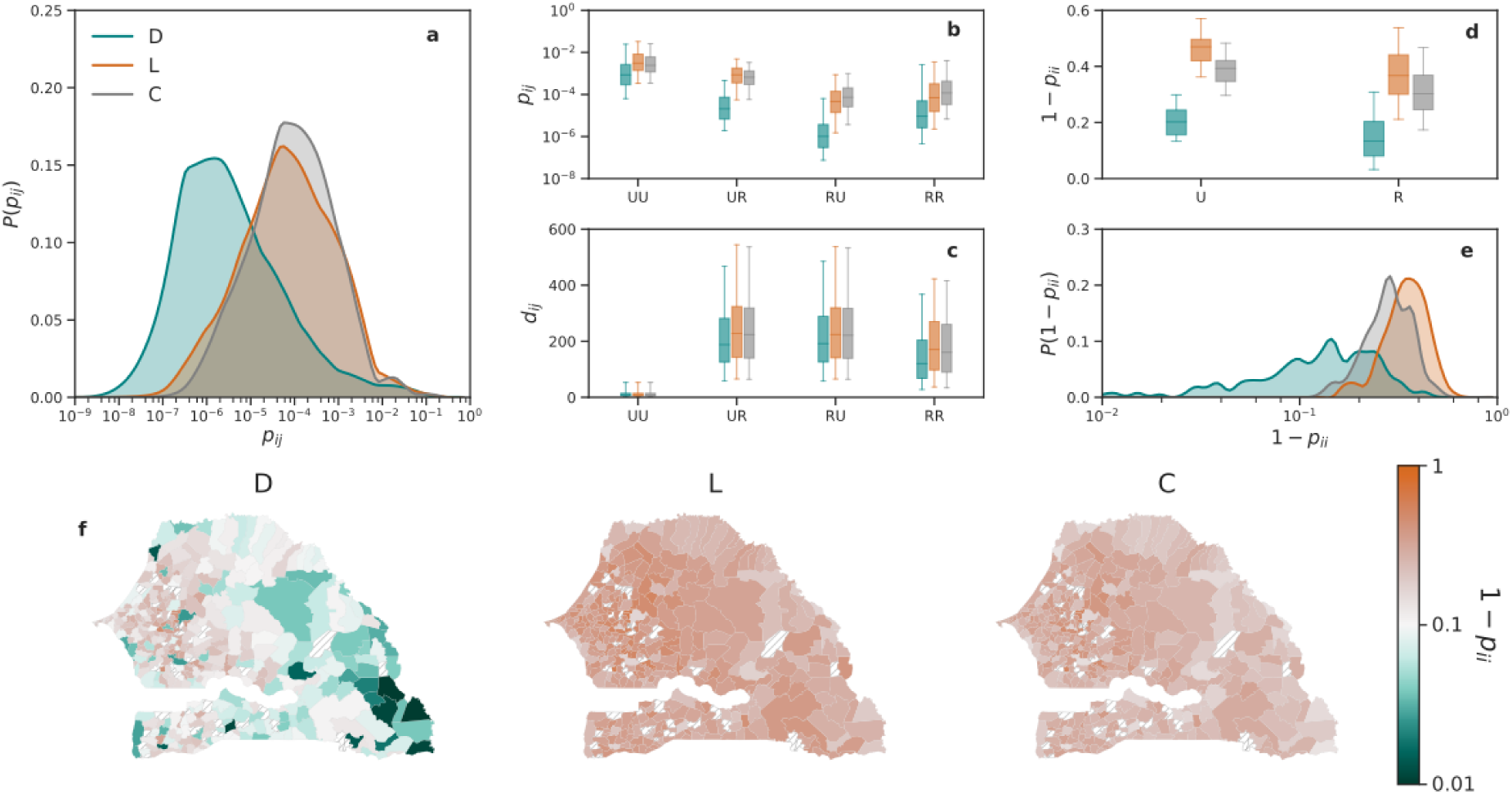
Coupling forces. a) Coupling probability distribution in *D*, *L*, *C*. Curves represent the average of the coupling probabilities among the 12 matrices. b),c),d) Box plots indicate the 95% reference range in January of the coupling probability, the outgoing probability distribution and the geographical distance, respectively. These represent a subset of links extracted by breaking down municipalities into Urban (U) and Rural (R). e) Outgoing probability distribution in *D*, *L*, *C*. f) Map of the outgoing probability in any municipality for *D*, *L* and *C*. Same distribution patterns have been found throughout thee years.

**Figure 3.**
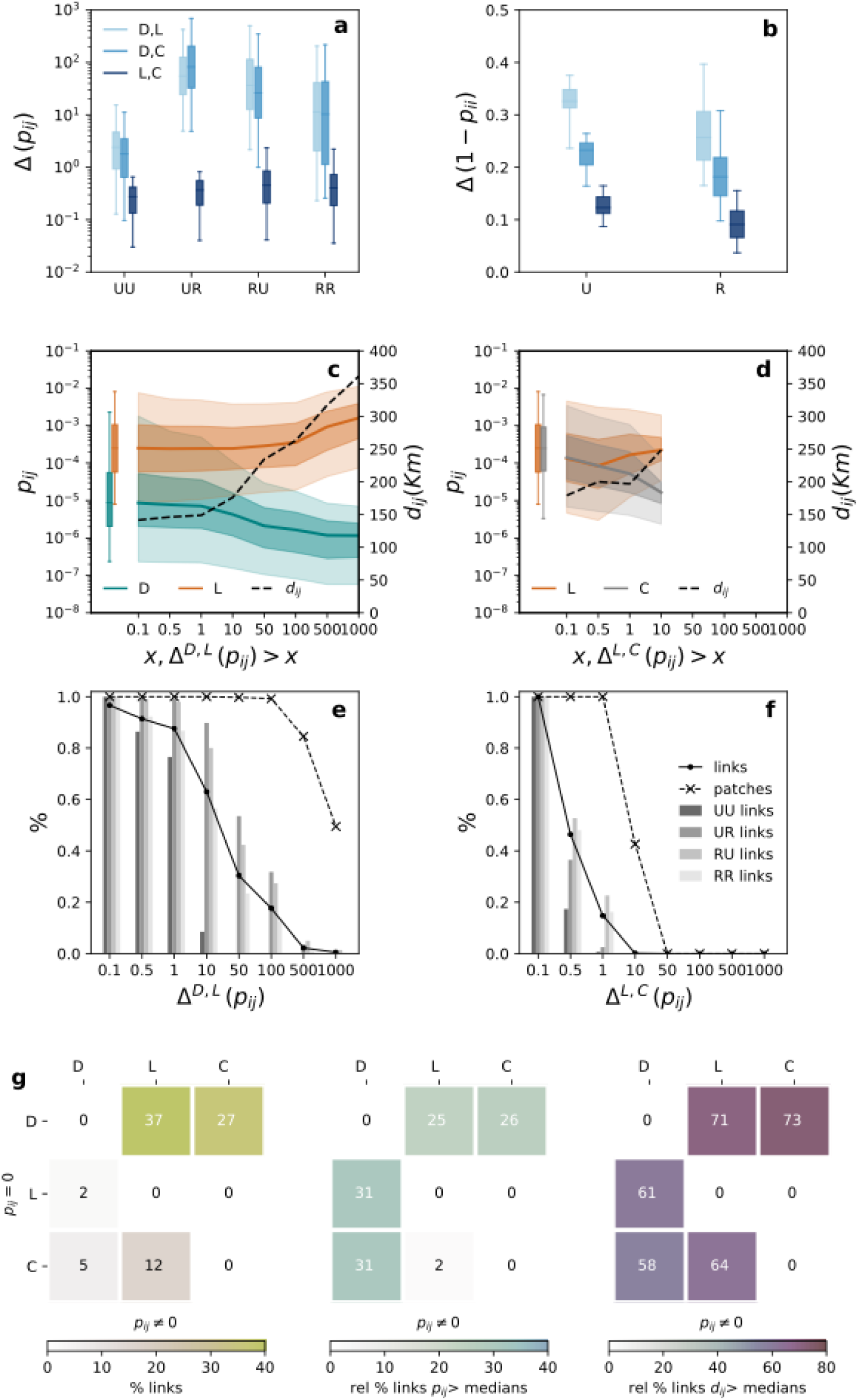
Differences between *D*, *L* and *C*. Box plots indicate the 95% reference range of the distribution of the relative difference of coupling probabilities of the common links among *D*, *L*, *C* and of the relative difference of outgoing probability of the common links among *D*, *L*, *C* in b). In c) and d) The curves display the coupling probability distribution for a subset of common links. Subsets of links are extracted a different cut-off of the relative variation between *D*, *L* and *L*, *C* on the coupling probability. The dotted lines show the median of the geographical distance distribution for the same subset of links. e) and f) show the percentage of links and patches present in any subset and the relative percentage of links extracted by breaking down municipalities into Urban (U) and Rural (R). g) Not-common links analysis. In the three matrices are shown the percentage of links detected by the aggregation method in the column and not detected by the aggregation method in the row (yellow matrix), the relative percentage of such not-common links with coupling probability higher than the median (green matrix) and the relative percentage of such not-common links with geographical distance higher than the median (violet matrix).

We focused the comparison of the three matrices on i) common links and ii) links detected in one method and not in another one.

i). In a) and b) we reported the relative variation distribution of the common links between any pair of methods. Considering subsets of links that have relative variation higher than a certain cut-off, we found that the biggest differences are between *D* and *L*, among links with the highest weight in *L* and the lowest in *D*. The probability of coupling in *D* can be up to 1000 times lower compared with the one in *L* (**Figure 3c,e**). We found these differences increase with the increasing of the geographical distance between the coupled municipalities (**Figure 3c**). In Methods is shown how we evaluated the geographical distance between municipalities. Same results have been found in the comparison between *D* and *C*. Instead, *C* and *L* are quite similar. We found links in *C* are no more than 10 times bigger than in *L* and their discrepancies are quite stable on the geographical distance. (**Figure 3d,f**).

ii). As **Figure 3** shows, we found that the 37% of the links in *L* (27% in *C*) are not detected in *D*, among those 25% of the links have a weight that is higher than the median (26% in C) (**Figure 3g**), and 71% (73% in *C*) are links between municipalities at long-range distance (**Figure 3g**). Instead, 12% of links detected in *L* do not exist in *C*. These links connect home locations to destinations in which people do secondary activities (See Method), presenting coupling probability lower than the median of coupling probability in *L* (**Figure 3**).

### Epidemic simulations

To assess the role of the aggregation approach on disease diffusion, we integrated the estimated coupling matrices in a spatially explicit metapopulation model. We considered three different epidemic scenarios: i) the top value in the confidential interval of Ebola Virus, *R*_0_ = 3 (high transmissibility); ii) the top value in the confidential interval of Influenza, *R*_0_ = 1.5 (medium transmissibility); iii) a scenario Influenza-like with control measures in action, *R*_0_ = 1.1 (low transmissibility).

For each scenario, we compared the output of the simulated epidemic equally initialized in *D*, *L*, *C*. We initialized the epidemic with 10 infected people and, we performed the simulations for 92 epidemic seeds (46 urban area and 46 rural one). Seeds are selected by considering all the urban areas and the top 10% of rural locations with the highest variation on outgoing and coupling probability between *D* and *L*. We investigated a set of global epidemics observables to characterize the simulated epidemic in time and space. We then based the comparison on two main observables: i) the arrival time of the infection in each location and ii) the invasion epidemic paths from the seed.

1. The arrival time. Over all, the lower coupling probability measured by *D* results in delayed arrival times. The arrival time in *D* ranges from few weeks to almost 410 days, while in *L* and *C* is quite lower, not exceeding 250 days. Arrival time in *D* and *C* are in accord to observations on spatial transmission driven by commuting flows modelled by^9^ with a similar metapopulation approach, but with different source to infer coupling forces. Considering *L* and *C*, the median of the relative variation on the arrival times is 0 for each *R*_0_ (**Figure 4a**) while it is quite higher (around 100%) comparing the two methods with *D*. We looked at the time when the 5% of the patches have been infected (*t*_5%_). As shown in Supplementary Information (**Figure 4**), the relative variation on *t*_5%_ between *L* and *C* do not exceed 200% and the median is 0, while between *D*,*L* and *D*,*C* it ranges from −100% to 1200% with a median of around 100%. In addition, *L* and *C* present a high correlation in the ranking of the arrival time in any location (Kendall tau coefficient ranging from 0.65 to 0.90). Kendall tau coefficient is not so high instead between *D* and the other two methods, **Figure 4b**. It means that *L* and *C* not only reproduce a quite similar distribution of the arrival time, but they also simulate the arrival of the epidemic in any place with the same ranking. Furthermore, the maximum geographical distance achieved at *t*_5%_ is much smaller in *D* compared with *L* and *C* (**Figure 4c**). As the map shows in **Figure 4d** when *R*_0_ = 1.1 the first 5% of municipalities infected in *D* are clustered close to the capital Dakar, while in *L* and C the epidemic invasion is more heterogeneous in space. Similar results with *R*_0_ = 1.5, and *R*_0_ = 3 are reported in the Supplementary Information.
2. The spatial invasion. We found *L* and *C* reproduce also similar heterogeneous paths of invasion in the country, accordingly with observed epidemic patterns of spatial invasion for emerging epidemics including H1N1 pandemic^35^, and the recent COVID-19 pandemic^36^. We selected only two locations as epidemic seed, Dakar and a rural area in the department of Saraya. The rural area is the furthest municipality from Dakar, and it is in the top 2% of municipalities with the highest variation in the outgoing probability between *D* and *L*. We computed the invasion trees by considering the likely epidemic invasion paths. We analyzed the similarity between invasion trees with a distance metrics based on the betweenness centrality (see Method). We found that the betweenness distance on the trees between *C* and *L* is lower compared with *D*,*L* and *D*,*C* as shown in **Figure 5** (*D,L* <0.05, *D*,*C* and *D*, *L* range from 0.05 to 0.25). Trees are shown in **Figure 5d**. We quantified the epidemic invasion distance (*d_inv_*) on the invasion trees as the number of hops from the seed needed to infect a given place (See Methods). We found that in *D* the invasion is mainly fragmented into short-distance hops (**Figure 5b,c**). Focusing on the first layer of infection (*d_inv_* = 1), as shown in **Figure 5d**, the infected locations are clustered close to the seed, while in *L* and *C* these are more heterogeneously distributed in space. It means that *C* and *L* reproduce more realistic epidemic patterns, integrating long-range transmission with local-range dispersal.

**Figure 4.**
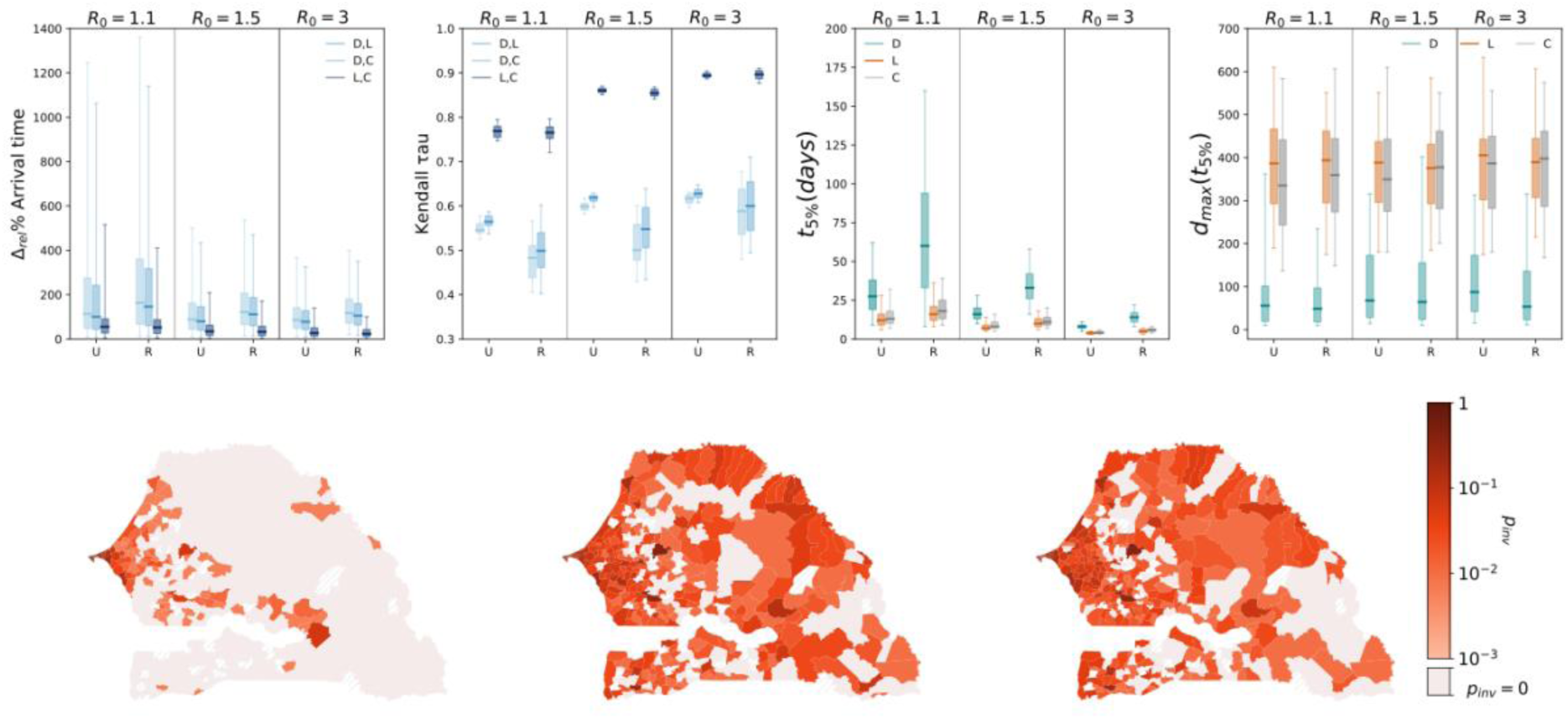
Differences on mode led epidemics. Box plots indicate the 95% reference range of a) The relative variation of the arrival times in any municipality between any pair of the three methods b) The Kendall tau probability by comparing *D*, *L*, *D*, *C* and *L*, *C* c) The maximum geographical distance achieved from the seed at *t*_5\%_. *t*_5\%_ is the time when the 5% of the municipalities have been infected (*t*_5\%_). Analysis of urban (U) and rural (R) epidemic seeds were done separately. d) Visualization on the map of the invasion probability *p_inv_* at *t*_5\%_ in the three methods with a municipality of the capital Dakar as seed and *R*_0_ = 1.1. The location with the red dot is the epidemic seed.

**Figure 5.**
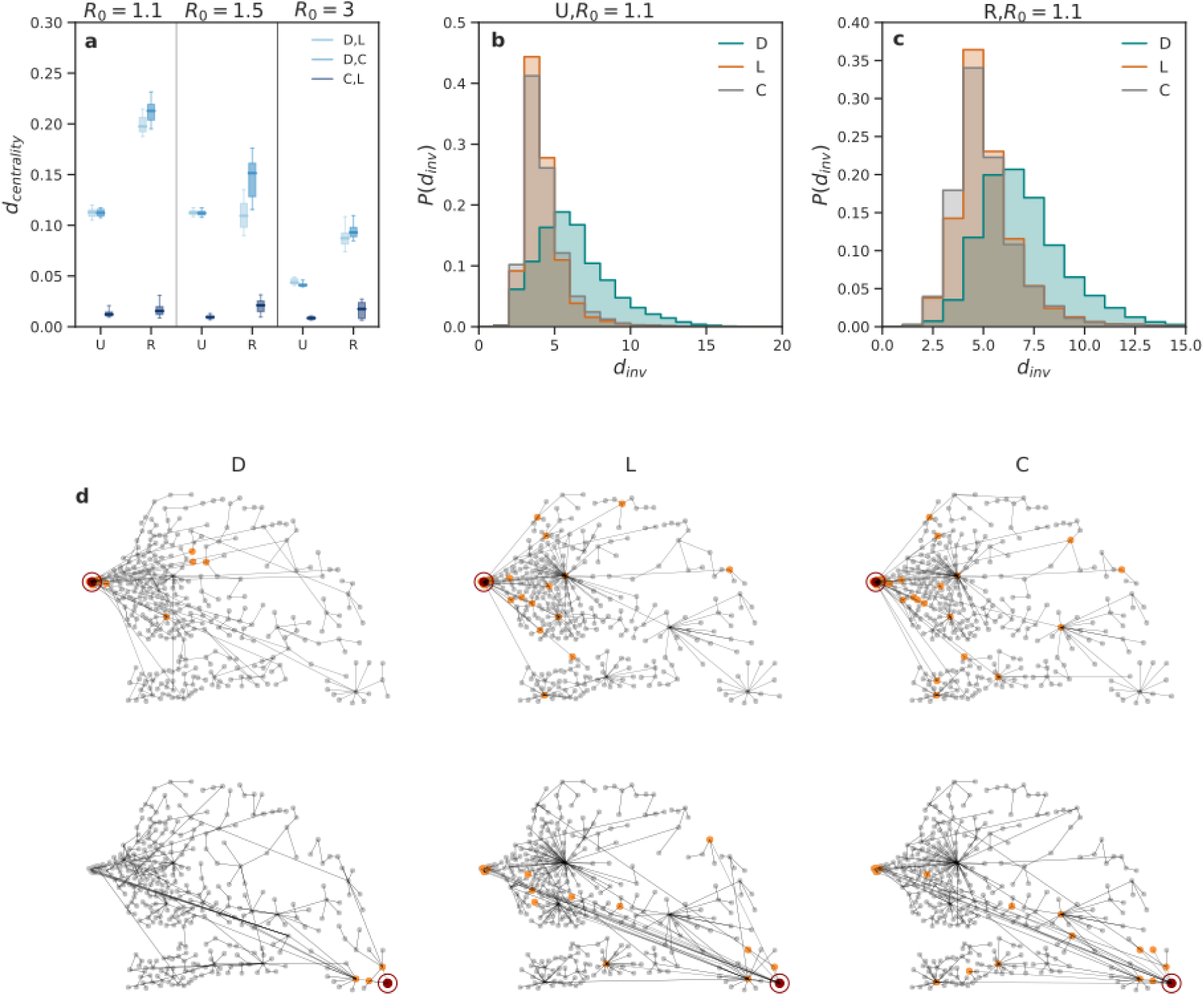
Epidemic invasion trees. Epidemic invasion trees have been computed for *R*_0_ = 1.1,1.5,3 considering both an urban (U) and a rural seed (R). a) Box plots indicate the 95% reference range of the betweenness centrality distance index measured between the epidemic infection trees of any pair of methods. b) and c) The distribution of the invasion distance in the three methods when *R*_0_ = 1.1. d) and e) The invasion trees when *R*_0_ = 1.1. Epidemic seeds are in red. Locations directly infected by the seed are in orange, while locations infected by other nodes are in gray.

Looking at the invasion trees in *D*, C and *L*, a key difference is played by the holy city Touba, the second most populated Senegalese city after the capital Dakar. For each method, Touba has always the role of the top spreader (i.e. maximum out-degree on the tree 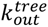. For instance, with *R*_0_ = 1.1, 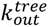 ranges from 11 to 19 in *D* and from 47 to 64 in *L* and *C*. In *C* and *L* around the 15% of the municipalities are infected by Touba, while in *D* less than 5%. In **Figure 4d** is shown that the invasion probability at *t*_5%_ in Touba in *L* and *C* is around one order of magnitude higher compared with *D*. Given the strong daily connection for commuting and commercial exchanges between Dakar and Touba^37^, results suggest that *L* and *C* better perform the modelled epidemics by detecting Touba as a relevant epidemic hotspot. This was also confirmed by the invasion dynamics of COVID-19 in Senegal which begun in Dakar and in few days reached also Toubap^38^. In any case, the differences between the simulated epidemic outcomes decrease, as expected, with high values of transmissibility (See SI).

Finally, we introduced the matrices *D’*,*L’*,*C’* to improves the previous matrices *D*,*L*,*C* and we analyzed the differences on the relative epidemic outcomes. We computed *L’* by considering the time spent in a place, interpolating exactly the time spent in any location based on the time slot of two consecutive calls, and not the number of calls as in *L*. *C’* is computed accounting for the most visited location over 12h (7am-7pm), and not over all 24h as in *C.* Reducing the time window, we aimed to capture the workplaces as most visited location. Last, *D’* is computed by considering the time elapse between any two displacement instead of the number of displacement as in *D*. We also defined *D_norm_* to try to well normalized *D* on the different mobile phone users’ activity profiles (See Supplementary Information).

Little differences exist between L, *L’* and *C*, *C’*, while relevant ones are found between *D* and *D’.* We found that in *L’* the outgoing probability decreases compared with *L*. It is probably a bias on the overestimation of the outgoing probability in *L* as users make few calls during the night and in this scenario the number of calls it is not a good proxy of time spent there. Relevant differences between *D* and *D’* could mean that users do a lot of consecutive calls in a short period in the same place, leading to an underestimation of the outgoing probability in *D*. Then, we found that the outgoing probability in *C’* is higher compared with *C* (**Figure 6a**). Intuitively, it means that the most visited location computed over the 12 daily hours more likely not match with the home location.

**Figure 6.**
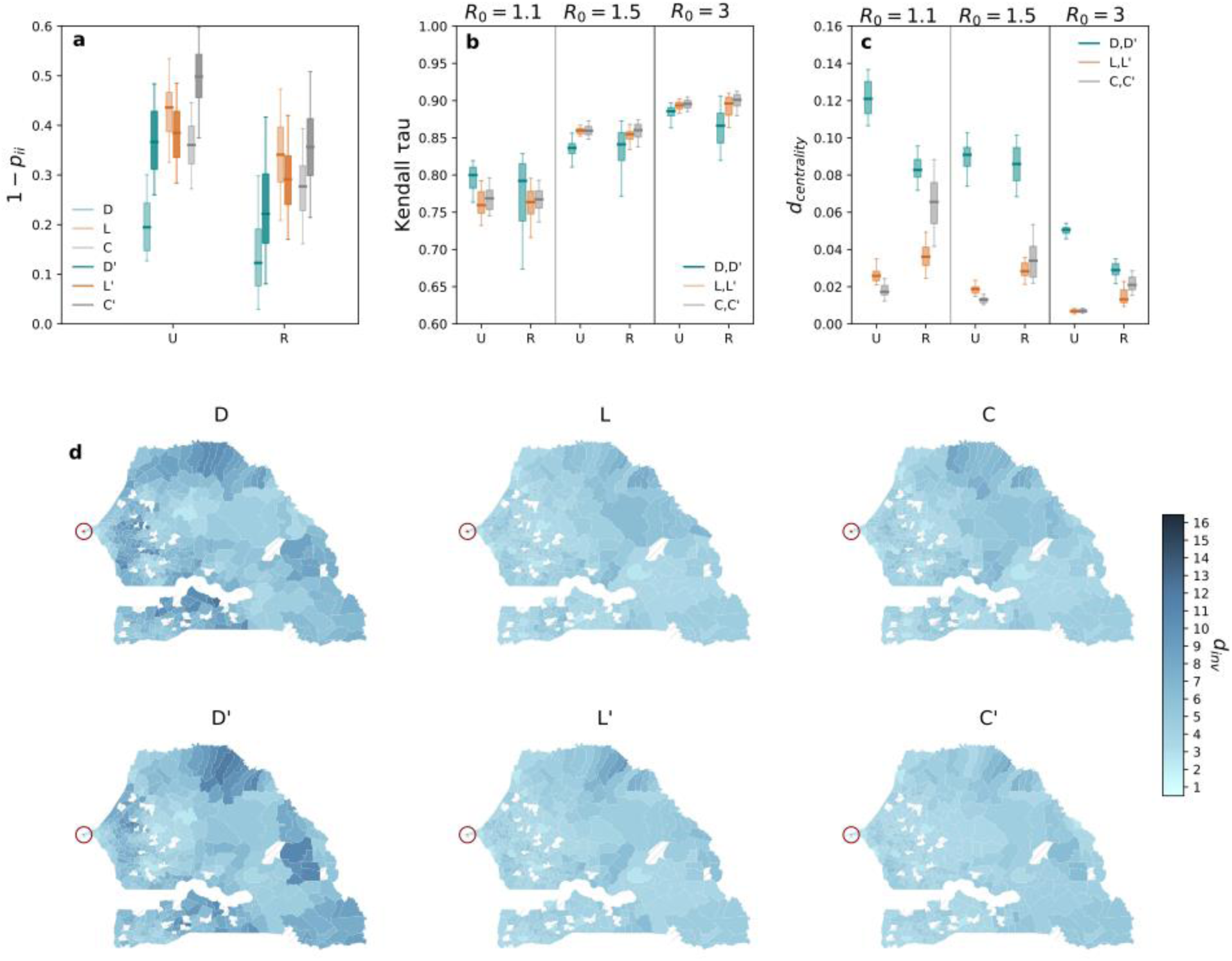
Sensitivity. a) The outgoing probability in January in *D*, *L*, *C* and *D’*, *L’*, *C’* by breaking down municipalities into Urban (U) and Rural (R) seeds, b) Kendall tau probability by comparing *D*, *D’*, *L*, *L’* and *C*, *C’* by breaking down into urban and rural seeds. c) Betweenness centrality distance index measured between the epidemic invasion trees of any pair of methods. Box plots indicate the 95% reference range. Betweenness centrality is measured on the invasion trees by selecting only an urban (U) and a rural seed (R). d) Visualization on map of the epidemic invasion distance when the capital Dakar (red circle) is the epidemic seed and *R*_0_ = 1.1.

Assessing the impact on the modelled epidemics, we have found that *L’* and *C’* do not involve crucial differences compared with *L* and *C* (**Figure 6b,c, d**). Meanwhile, *D’* reproduced a spatial diffusion that is more similar to *L* and *C*. However, the invasion distance distribution in *D’* is higher compared with *L* and *C*. It depends on the 40% of links missing in *D’* existing in *L* and *C*. It suggests that *D* and *D’* are not well-defined to detect epidemiological couplings among locations.

Our findings indicate that preserving the full resolution of the observed trajectory of individual movements (*D*,*D’*) may bias the spatio-temporal diffusion of the simulated epidemic in both the timing and pattern of invasion. Instead, the aggregation of visited locations (*L*,*L’*), while loosing all information about their sequence in an individual path, reproduce realistic simulated patterns. High similarity between *L*,*L’* and *C*,*C’* suggests that for a range of epidemic contexts, secondary activities have no significant impact on the spread. While, places in which individuals spent most of their time are the main driver of disease diffusion, as it was found in the largest US metropolitan areas in which researchers found that the majority of COVID-19 infections came in a subset of highly visited points of interests^27^. *D* and *D’* could be a good option in the case is important to know the actual path of the individuals, such as migration processes, in which it is important to track displacements from previous resident place to new home location.

## Discussion

In the last decades, mobile phone data have been largely incorporated into epidemic models to study how epidemics transmission occur^6^. Mobile phone traces indeed have been helpful to understand the spatial transmission of many outbreaks including malaria^13,15^, cholera^12,16^, schizosomiasis^17^, Ebola^21,22^, dengue^18^, HIV^19,20^.

Since the beginning of COVID-19 pandemic, mobile phone data has been used to help fight the public health emergency. In this context, many network operators and private companies made a huge effort to rapidly share their data under legally and ethically compliant frameworks^25,26,33^. Researchers all over the world started dealing with such high-resolution data to track human behavior and inform epidemic models^23,27–29^.

Adequately aggregating human movements become particularly relevant for improving reliability of projections of infection diseases models. On the other hand, it is also become crucial in the assessment of the meaningful mobility information needed to inform models in order to minimize the sharing of sensitive data between data providers and researchers.

To answer this questions, we extracted three coupling matrices, *D*,*L*,*C* at higher, medium and lower resolution, respectively, and we aimed to assess the different impacts of using *D*, *L* and *C* to simulate emerging epidemics. We designed an implicit metapopulation model. The matrices constituted the input data to inform our spatial modelling framework. We proposed a new non-Markovian model in which the force of infection in a given place is based on three categories of infected people: people not moving, visitors, returning residents. In the model, we introduced a normalization factor on the effective population that is composed of all these three categories of people.

We found two key results: i) preserving maximum resolution of the individual trajectories involves a bias in the simulated epidemics; ii) the only information on the daily users’ most visited location is sufficient to model the spatial transmission. Keeping the full resolution of the individual trajectories, Displacement-based method (*D*) involves a delay in the simulated epidemics. Accounting for any displacement of the users, in *D* the epidemic invasion is mainly fragmented in short-range hops, and it presents a radial diffusion.

Instead, *C* and *L* not consider the actual trajectory, but they take into account the home of the individuals and the time spent in each location. This allows to account for the duration of travels, which plays a crucial role in shaping epidemics^39,40^. This approach capture the locations in which individuals spend time and the ones in which they only pass by. Due to that, *L* and *C* reproduce more realistic spatial and temporal epidemic patterns by simulating both short than long-range contagions. The high similarity between *L* and *C* suggests that all secondary activities considered in *C* are less relevant in the infection dynamics.

We underlined the limitations to use *D* approach. *D* by fragmenting the actual trips in several displacements does not capture the origin and destination locations. A possible solution could be to correct this bias by identifying the anchor points of people’s call activities, and then defines movements between different anchor points as trips as in ^41^. Moreover, *p_ij_* in *D* is defined accounting for users’ displacements at different time scales, involving a limitation at defining the timescale of the simulations in modeling. Also *D_norm_* which is properly normalized over the different user profiles, however, not reproduce realistic epidemics patterns. It suggests that the Displacement-based approach could not be used to model emerging epidemics with a metapopulation approach. This approach might be instead considered modelling radial diffusion, such as to model epidemic invasion due to migrations flows.

We are aware that this work has some limitations. Mobile phone’s access and usage depend on demographic and socioeconomic properties of individuals. In most countries, mobile phone billing is proportional to the number of call and SMS, generating a large inequality in individuals’ activity and thus in data representativeness between social strata^42^. Data is indeed most representative of urban populations for target subgroups, suffering from demographic and geographical representativeness, and other issues concerning ownership, mobile phones sharing and, heterogeneity in cell tower distribution and in individuals’^42–45^.

However, the aggregation of mobile phone individual trajectories into coupling forces thought a metapopulation approach allows to partially avoid mobile phone biases^46–48^. To reduce biases, we aggregated individual trajectories at cell tower level to coupling forces at municipality level, breaking down locations in urban and rural areas. I also proposed the aggregation approach *L’* and *D’* to correct the heterogeneity in individuals’ activity. Moreover, the metapopulation structure is designed at municipality level, averaging the time spent into indoor and outdoor locations. We are aware transmissibility in these places are different, however, our aim was to compare modelled outcomes by varying mobility definition and resolution at global level to reproduce epidemic spatial invasion, and not to design an ad hoc model for specif locations.

Further work will be focus on comparing mobile phone aggregation procedures at different spatial scales, aiming to assess if our results are robust across the spatial granularity of the metapopulation structure. However, reducing the spatial scale means averaging on all the heterogeneities, so we expect the differences between the aggregation methods will drastically decrease. Our study was performed in Senegal in a peacetime period, i.e. ongoing epidemics were not present. We expect results will not change in other developing countries having a similar cultural, social, and economic situation. Last, the work is not validated on epidemiological data. However, the heterogeneous invasion patterns modelled in *L* and *C* has been observed during the COVID-19 epidemic in Senegalp^38^. In other countries, heterogeneous invasion patterns have been also experienced both for epidemics with a medium transmissibility rate like Influenza^49^ and in faster epidemics like Ebola^50^. Further investigations are, however, needed to validate the results on epidemiological data.

## Methods

### Dataset description

The research is based on the analysis of pseudo-anonymised mobile phone data collected by Orange. The dataset consists in a set of Call Detail Records (CDRs) of phone calls and text exchanges between Orange’s customers generated every time that a person does an activity by making a call or sending a message. Each record contains several attributes: the caller and callee IDs, the time-stamp, the duration of the activity, the type of communication (national, international, call outgoing, call incoming) and the identifier of the antenna that handled the activity. We analyzed a set of 15,859,942,126 records of 9,569,425 million mobile phone users (80 % of the country population) from January to December 2013 handled by 15999 Orange antennas. The antennas are heterogeneously distributed over the whole territory of Senegal, covering all 46 urban municipalities and 357 out of 437 rural ones. Given the fluctuations in time of the number of active antennas, the number of covered municipalities is time-varying over the year. So, we considered only the municipalities covered throughout the entire 2013. We thus selected 394 ones (46 urban and 348 rural). We retained only users who have been active more than 30 days and with less than 1000 mobile activities per week. Since shared phones is a phenomenon very common in Africa^44^, the threshold on the maximum number of activities per week allows avoiding bias due to multi-ownership of the phone.

### Coupling Matrices

Mobile phone data allows tracking individual daily trajectories by interpolating every user’s displacements based on two consecutive activities (calls/SMSs). By aggregating in time and space trajectories of all users, it can be defined a matrix, so-called Coupling Matrix, for estimating the probability of epidemiological coupling between locations. We quantified the impact of different levels of aggregation by extracting three coupling matrices largely used in literature at highest, medium and lowest resolution^11,12,44^. For each method, we extracted 12 monthly coupling matrices at municipality level. The coupling matrices of the three methods as defined as follows:

Displacement-based coupling matrix *D*

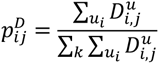

where 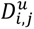 is the number of times user *u* moves from location *i* to *j*. This method is at high resolution as it keeps the temporal sequence of each displacement of the users. The coupling probability between two locations is defined as the probability of moving from one place to another. In this approach, the time spent by users in any place is not considered. In fact, each displacement has the same weight in the total sum, regardless of the time elapsed between two consecutive displacements. In a further work^51^, Lima et al. introduced another formulation where 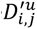 is re-defined as the sum of the time elapsed between any displacement between *i* and *j* done by the user *u*. We called this new approach *D*′. The new definition aimed to correct *D* by reducing the bias due to the heterogeneity in time and space of the number of calls made by mobile phone users. To improve *D*, we defined also *D_norm_*, where 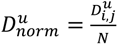 where *N* is the number of activities of the user u. Location-based coupling matrix *L*

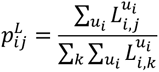

where 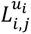 is the number of calls made in *j* by user *u_i_* living in *i*. Users living in *i* are detected by extracting individuals that do the most number of calls in *i* during nighttime - from 7 pm to 7 am. This is a well-established method used in literature to define the home location of mobile phone users^7^. In this method, the information of the temporal sequence of any individual displacement is lost, and the coupling is proportional to the number of calls made by users in each location. By assuming that the number of calls made by users in a place is proportional to the time that they spend in it, this method measures the coupling between any two locations *i-j* as the probability of being in j living in *i.* In this approach, all the locations users visits are divided into places where they spend most of their time and places where they just spend few minutes. However, by accounting for the number of calls, in this case as in the displacement-based one, there is a bias due to the heterogeneity of the activity of mobile phone users.

To improve *L*, we defined also 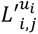 where 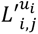 is the time spent in *j* by a user *u_i_* living in *i*. We extracted the effective time spent in a given location by looking at the time elapsed between two consecutive calls and assuming users spent half of the time in the origin location, and half of the time in the destination location.

Most visited location-based coupling matrix *C*

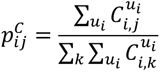

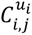 is the amount of time user *u_i_* spends at his most visited location daily in *j*. The most visited location is measured as the location where a user makes the maximum number of calls during all day (24h). Therefore, this method takes only locations where users spend most of their time into account (home, work or school places). Any other less visited location is neglected. As in the Location based one, the coupling between any two locations *i-j* in this method does not represent the actual probability of moving from *i* to *j.* In this case the coupling probability is the probability of spending most of the time in *j* living in *i.* Since the most visited location often coincides with the work location of an individual, this aggregation process may be considered an extraction of the commuting fluxes. Tizzoni et al. ^9^ found similar patterns in three European countries.

To improve C, we defined also 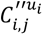, where 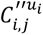, is the number of time that user *u*_i_ has his/her daily most visited location in *j*. In *C*’, the most visited location is measured as the location where a user makes the maximum number of calls during the daytime (7am-7pm).

### Epidemic metapopulation model

To quantify the impact of the coupling matrices on the modelled epidemic diffusion, we computed numerical simulations based on a metapopulation approach. We developed a new stochastic, discrete and Non-Markovian model. It accounts for disease transmission due to visitors and to staying and returning residents. In fact, people leaving in a location are exposed to an infection carry by (i) people not moving (ii) visitors and (iii) residents infected in other locations. The Senegalese population is spatially divided in 394 municipalities depending on census data and links between municipalities are defined from coupling matrices. We considered a SEIR (Susceptible - Exposed - Infectious - Recovered) epidemic dynamics in a closed population with no births or deaths^52^. The discrete-time SEIR model has the following form:

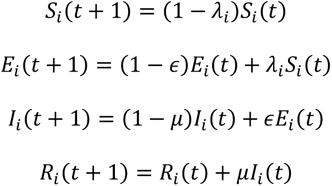

*λ_i_*, *∈*, *μ* are the force of infection in a subpopulation *i*, the incubation rate and the recovery rate respectively. *S_i_*(*t*), *E_i_*(*t*), *I_i_*(*t*), *R_i_*(*t*), denote the number of susceptible, exposed, infected and recovered individuals at time *t*. For every *i*, *N_i_* = *S_i_* + *E_i_* + *I_i_* + *R_i_* where *N_i_* is the number of resident in the municipality *i*.

Taking into account the coupling between patches, the force of infection in a node *i* is calculated as:

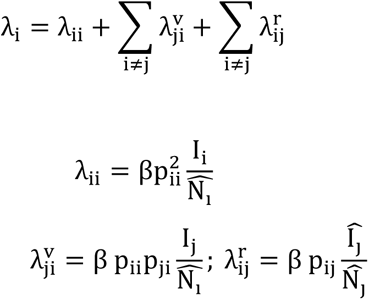

where *p_ij_* is the coupling probability between patches *i* and *j*,

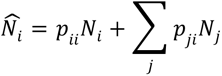

is the effective population in *i* and

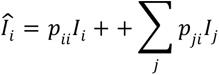

is the effective number of infections.

The initialization of the simulations consists in setting in one patch *i*=10 infected individuals. We set the parameters as follows: the average incubation period is *∈^−^*^1^ = 1.5 days, the average duration of the infection is *μ^−^*^1^ = 3days. We explored different values of the basic reproduction number *R*_0_ = 1.1,1,5,3, low, medium, high transmissibility, respectively. Stochastic simulations are computed by assuming 1 day as time step of the simulation.

### Analysis

#### Comparison between coupling matrices

To underline similarities and differences of the three matrices, we compared any pairs of associated coupling networks. We analyzed separately two subsets of links, i) links detected in both the two compared matrices, ii) links detected only in one of the two compared matrices or links not detected in both. In i) we performed a network regression analysis (MRQAP) to test the correlation between coupling probability of the links for each pair of matrices^53^. We explored how relative variation among matrices of coupling probabilities are related or not with the geographical distance. Then, to study the correlations of coupling patterns among the 12-month time periods of the matrices, we implemented a hierarchical clustering. For each month, we constructed the vector of all pairwise coupling probability among the locations. Then, we implemented the Pearson correlations *ρ* among any two vectors of the 12 monthly periods, and we define a dissimilarity matrix 12 x 12 in which each element *m_ij_* = 1 *− ρ_ij_*. The hierarchical clustering is evaluated on the euclidean distance between any two element of the matrix. Instead, in ii) we computed the percentage of links in one matrix that are not detected in the others, and we put in relation the percentage of these not-detected links with the geographical distance among not-coupled locations. Geographical distance is computed between any two locations by using the Haversine Formula. Last, we looked at the outgoing probability in each municipality. The outgoing probability is defined as follows:

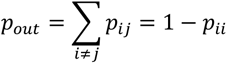

#### Comparison between simulated epidemic outcomes

By assessing how the level of the spatio-temporal aggregation of the individual trajectories impacts the modelled epidemic diffusion in space and in time, we kept track of the following epidemic observables. First, we investigated the temporal diffusion of the epidemic, measuring the arrival time t_a of the epidemic in any location. The arrival time t_a of the epidemic is the first time that an individual become infected in a given municipality. The probability distribution of the arrival time and its median value are evaluated for every location in each synthetic scenario. We also evaluated the Kendall tau correlation coefficient for exploring if the ranking of the epidemic arrival time. By tracking the spatial diffusion of the simulated epidemic, we measured the epidemic invasion tree^2,9^ which represents the most likely transmission path of the infection over all the duration of the epidemic. Considering a disease-free location *i*, as soon as *I_i_ ≠* 0, we tracked a directed link between *i* and the location of origin of the infected individual. For each scenario, for each run, we extracted the invasion path. Then, by cumulating over the runs, we obtained a unique invasion path where the weight of the links is the percentage of the number of runs in which that link exists. Once the invasion path is obtained, we calculated the invasion tree by measuring the direct maximum spanning tree. To avoid the stochastic fluctuations of the epidemic simulations, we computed invasion tress by doing 1000 runs, and selecting randomly for 50 times 400 runs. We compared the invasion trees through two metrics: the betweenness centrality distance^54^ and the invasion distance. The betweenness centrality distance between coupling matrices *X* and *Y* is defined as follows:

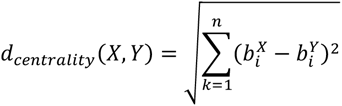

where the betweenness centrality of a node *i* is given by the expression:

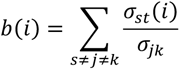

*σ_jk_* is the total number of the shortest paths from node *j* to node *k* and *σ_jk_*(*i*) is the number of those paths that pass through *i.* The invasion distance in any invasion tree is measured for each node, and it is the number of edges connecting the considered node with the root. We also computed the invasion probability in any node, considering both urban and rural seeds. We defined the invasion probability *p*(*t*)*_inv_* for a patch *i* as the probability that the epidemic reaches *i* in a given time *t*.

## Data Availability

Mobile phone data are proprietary and confidential. For legal reasons, the full dataset is not publicly available. However, access to the aggregated data can be requested from Orange authors on a contractual basis.

## Acknowledgments

This study was partially funded by: Agence Nationale de la Recherche project DATAREDUX (ANR-19-CE46-0008-03); Horizon Europe grants ESCAPE (101095619) and VERDI (101045989); EU Horizon 2020 grant MOOD (H2020-874850). The contents of this publication are the sole responsibility of the authors and don’t necessarily reflect the views of the European Commission.

## References

1. Colizza, V., Barrat, A., Barthélemy, M. & Vespignani, A. The role of the airline transportation network in the prediction and predictability of global epidemics. Proc. Natl. Acad. Sci. U. S. A. 103, 2015–2020 (2006).

2. Balcan, D. et al. Multiscale mobility networks and the spatial spreading of infectious diseases. Proc. Natl. Acad. Sci. 106, 21484–21489 (2009).

3. Colizza, V., Barrat, A., Barthelemy, M., Valleron, A.-J. & Vespignani, A. Modeling the worldwide spread of pandemic influenza: baseline case and containment interventions. PLoS Med. 4, e13 (2007).

4. Barbosa-Filho, H. et al. Human Mobility: Models and Applications. Phys. Rep. 734, 1–74 (2018).

5. Eurostat. https://ec.europa.eu/eurostat/.

6. Maxmen, A. Can tracking people through phone-call data improve lives? Nature 569, 614–617 (2019).

7. Blondel, V. D., Decuyper, A. & Krings, G. A survey of results on mobile phone datasets analysis. EPJ Data Sci. 4, 10 (2015).

8. The Mobile Economy. *The Mobile Economy* https://www.gsma.com/mobileeconomy/.

9. Tizzoni, M. et al. On the Use of Human Mobility Proxies for Modeling Epidemics. PLOS Comput. Biol. 10, e1003716 (2014).

10. Panigutti, C., Tizzoni, M., Bajardi, P., Smoreda, Z. & Colizza, V. Assessing the use of mobile phone data to describe recurrent mobility patterns in spatial epidemic models. R. Soc. Open Sci. 4, 160950 (2017).

11. Lima, A., Domenico, M. D., Pejovic, V. & Musolesi, M. Disease Containment Strategies based on Mobility and Information Dissemination. Sci. Rep. 5, 10650 (2015).

12. Finger, F. et al. Mobile phone data highlights the role of mass gatherings in the spreading of cholera outbreaks. Proc. Natl. Acad. Sci. U. S. A. 113, 6421–6426 (2016).

13. Wesolowski, A. et al. Quantifying the impact of human mobility on malaria. Science 338, 267–270 (2012).

14. Rubrichi, S., Smoreda, Z. & Musolesi, M. A comparison of spatial-based targeted disease mitigation strategies using mobile phone data. EPJ Data Sci. 7, 1–15 (2018).

15. Tatem, A. J. et al. Integrating rapid risk mapping and mobile phone call record data for strategic malaria elimination planning. Malar. J. 13, 52 (2014).

16. Bengtsson, L. et al. Using mobile phone data to predict the spatial spread of cholera. Sci. Rep. 5, 8923 (2015).

17. Ciddio, M. et al. The spatial spread of schistosomiasis: A multidimensional network model applied to Saint-Louis region, Senegal. Adv. Water Resour. 108, 406–415 (2017).

18. Wesolowski, A. et al. Impact of human mobility on the emergence of dengue epidemics in Pakistan. Proc. Natl. Acad. Sci. U. S. A. 112, 11887–11892 (2015).

19. Isdory, A., Mureithi, E. W. & Sumpter, D. J. T. The Impact of Human Mobility on HIV Transmission in Kenya. PLoS ONE 10, (2015).

20. Valdano, E., Okano, J. T., Colizza, V., Mitonga, H. K. & Blower, S. Using mobile phone data to reveal risk flow networks underlying the HIV epidemic in Namibia. Nat. Commun. 12, 2837 (2021).

21. Peak, C. et al. Population mobility reductions associated with travel restrictions during the Ebola epidemic in Sierra Leone: use of mobile phone data. Int. J. Epidemiol. 47, (2018).

22. Wesolowski, A. et al. Commentary: Containing the Ebola Outbreak – the Potential and Challenge of Mobile Network Dataeffe. PLOS Curr. Outbreaks (2014) doi:10.1371/currents.outbreaks.0177e7fcf52217b8b634376e2f3efc5e.

23. Pullano, G., Valdano, E., Scarpa, N., Rubrichi, S. & Colizza, V. Evaluating the effect of demographic factors, socioeconomic factors, and risk aversion on mobility during the COVID-19 epidemic in France under lockdown: a population-based study. *Lancet Digit*. Health 2, e638–e649 (2020).

24. Pullano, G. et al. Underdetection of cases of COVID-19 in France threatens epidemic control. Nature 590, 134–139 (2021).

25. Google.com. COVID-19 Community Mobility Report. COVID-19 Community Mobility Report https://www.google.com/covid19/mobility?hl=fr. https://www.google.com/covid19/mobility/.

26. Facebook Data for Good. *Facebook Data for Good* https://dataforgood.fb.com/.

27. Chang, S. et al. Mobility network models of COVID-19 explain inequities and inform reopening. Nature 589, 82–87 (2021).

28. Pepe, E. et al. COVID-19 outbreak response, a dataset to assess mobility changes in Italy following national lockdown. Sci. Data 7, (2020).

29. Gatto, M. et al. Spread and dynamics of the COVID-19 epidemic in Italy: Effects of emergency containment measures. Proc. Natl. Acad. Sci. 117, 10484–10491 (2020).

30. Bansal, S., Chowell, G., Simonsen, L., Vespignani, A. & Viboud, C. Big Data for Infectious Disease Surveillance and Modeling. J. Infect. Dis. 214, S375–S379 (2016).

31. Wesolowski, A., Buckee, C., Engø-Monsen, K. & Metcalf, C. J. Connecting Mobility to Infectious Diseases: The Promise and Limits of Mobile Phone Data. J. Infect. Dis. 214, S414–S420 (2016).

32. Montjoye, Y.-A., Hidalgo, C., Verleysen, M. & Blondel, V. Unique in the Crowd: The Privacy Bounds of Human Mobility. Sci. Rep. 3, 1376 (2013).

33. Oliver, N. et al. Mobile phone data for informing public health actions across the COVID-19 pandemic life cycle. Sci. Adv. 6, eabc0764 (2020).

34. Grantz, K. H. et al. The use of mobile phone data to inform analysis of COVID-19 pandemic epidemiology. Nat. Commun. 11, 4961 (2020).

35. Bajardi, P. et al. Human Mobility Networks, Travel Restrictions, and the Global Spread of 2009 H1N1 Pandemic. PLOS ONE 6, e16591 (2011).

36. Pinotti, F. et al. Tracing and analysis of 288 early SARS-CoV-2 infections outside China: A modeling study. PLOS Med. 17, e1003193 (2020).

37. Thiam, O. L’axe Dakar-Touba (Sénégal) : analyse spatiale d’un corridor urbain émergent. (2008).

38. Petit, V., Robin, N. & Martin, N. Spatialité et temporalité de l’épidémie de la Covid-19 au Sénégal. Le processus de production des données sanitaires au regard des discontinuités territoriales. Rev. Francoph. Sur Santé Territ. (2021) doi:10.4000/rfst.1150.

39. Poletto, C., Tizzoni, M. & Colizza, V. Human mobility and time spent at destination: Impact on spatial epidemic spreading. J. Theor. Biol. 338, 41–58 (2013).

40. Giles, J. R. et al. The duration of travel impacts the spatial dynamics of infectious diseases. Proc. Natl. Acad. Sci. 117, 22572–22579 (2020).

41. Kang, C., Sobolevsky, S., Liu, Y. & Ratti, C. Exploring human movements in Singapore: A comparative analysis based on mobile phone and taxicab usages. in vol. 1 (2013).

42. Schlosser, F., Sekara, V., Brockmann, D. & Garcia-Herranz, M. Biases in human mobility data impact epidemic modeling. ArXiv211212521 Phys. Q-Bio (2021).

43. Kishore, N. et al. Measuring mobility to monitor travel and physical distancing interventions: a common framework for mobile phone data analysis. *Lancet Digit*. Health 2, e622–e628 (2020).

44. Wesolowski, A., Eagle, N., Noor, A. M., Snow, R. W. & Buckee, C. O. Heterogeneous Mobile Phone Ownership and Usage Patterns in Kenya. PLoS ONE 7, (2012).

45. Chen, B. Y. et al. Understanding the Impacts of Human Mobility on Accessibility Using Massive Mobile Phone Tracking Data. Ann. Am. Assoc. Geogr. 108, 1115–1133 (2018).

46. Ma, K. C. & Lipsitch, M. Big data and simple models used to track the spread of COVID-19 in cities. Nature 589, 26–28 (2021).

47. Ajelli, M. et al. RCesoeamrchpartaicrleing large-scale computational approaches to epidemic modeling: Agent-based versus structured metapopulation models. 13 (2010).

48. Jia, J. S. et al. Population flow drives spatio-temporal distribution of COVID-19 in China. Nature 582, 389–394 (2020).

49. Coletti, P., Poletto, C., Turbelin, C., Blanchon, T. & Colizza, V. Shifting patterns of seasonal influenza epidemics. Sci. Rep. 8, (2018).

50. Kraemer, M. U. G. et al. Utilizing general human movement models to predict the spread of emerging infectious diseases in resource poor settings. Sci. Rep. 9, 5151 (2019).

51. Lima, A., Pejovic, V., Rossi, L., Musolesi, M. & Gonzalez, M. Progmosis: Evaluating Risky Individual Behavior During Epidemics Using Mobile Network Data. ArXiv150401316 Phys. (2015).

52. Keeling, M., Rohani, P. & Pourbohloul, B. Modeling Infectious Diseases in Humans and Animals. Clin. Infect. Dis. Off. Publ. Infect. Dis. Soc. Am. 47, 864–865 (2008).

53. Dekker, D., Krackhardt, D. & Snijders, T. A. B. Sensitivity of MRQAP Tests to Collinearity and Autocorrelation Conditions. Psychometrika 72, 563–581 (2007).

54. Donnat, C. & Holmes, S. Tracking network dynamics: a survey of distances and similarity metrics. ArXiv180107351 Phys. Stat (2018).

