## Supplementary Material for "Mobility resolution needed to inform predictive epidemic models for spatial transmission from mobile phone data"

### Supplementary Information

In the  Supplementary Information, we present the additional results of the work and the sensitivity analysis. More details are provided both on the statistical comparison of the coupling matrices and on the outcomes of the epidemic simulations.

Statistical comparison on coupling matrices

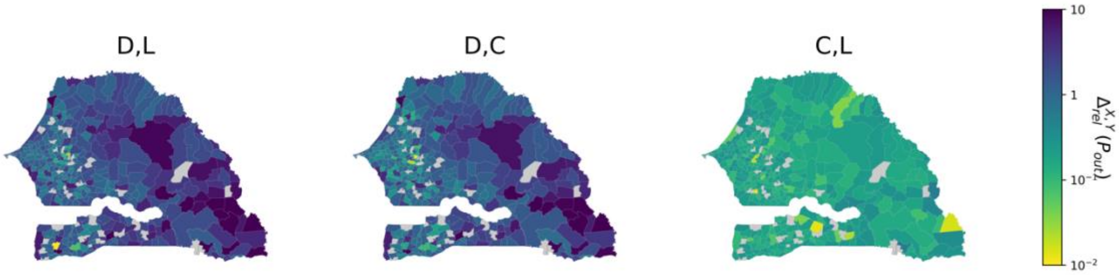


**Figure S1.** Relative variation of the outgoing probability between *D*,*L*, *D*,*C* and *C*,*L*.

**Figure S1.** shows that the height differences are between *D*, *L* and *D*, *C.* The relative variation of the outgoing probabilities  between D and *L*, *C* ranges from 1 to 10 except few outliers, while the relative variation between *L* and *C* does not exceed 1.

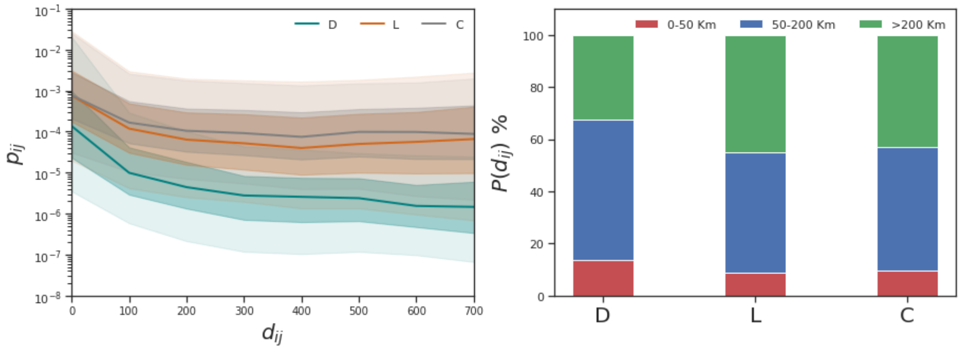

**Figure S2a**. a) Relation between the coupling probability and the geographical distance of the connected links in *D*, *L*, *C*. b) Percentage of the connected municipalities in *D*, *L*, *C* in a given geographical distance range.

Studying the relation between the coupling probabilities and the geographical distance of the connected links, in **Figure S2a** is shown, as expected, that the coupling probability decreases with the decreasing of the geographical distance in any method. In **Figure S2b** is shown that in *D* the percentage of links connecting municipalities longer than 200Km is around 70%, while in *L* and *C* is around 50%.

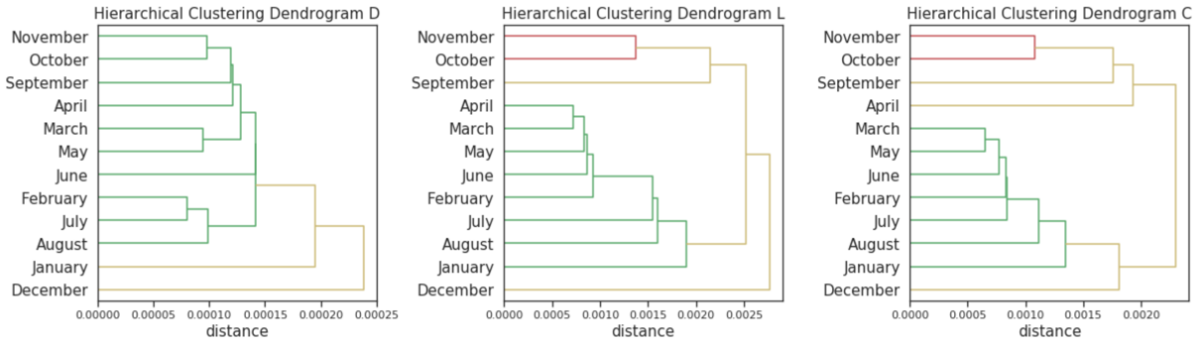


**Figure S3.** Hierarchical clustering Dendogram in *D*, *L*, *C*. Colors represent the resulting clusters between all months.

Moreover, we implemented a hierarchical clustering (See Methods) to study the correlations of the coupling patterns of the matrices among the 12-month time period. In **Figure S3** is shown how *L* and *C* reproduce quite the same clusters over the years; while they were not assessed in *D.*

Epidemic simulations

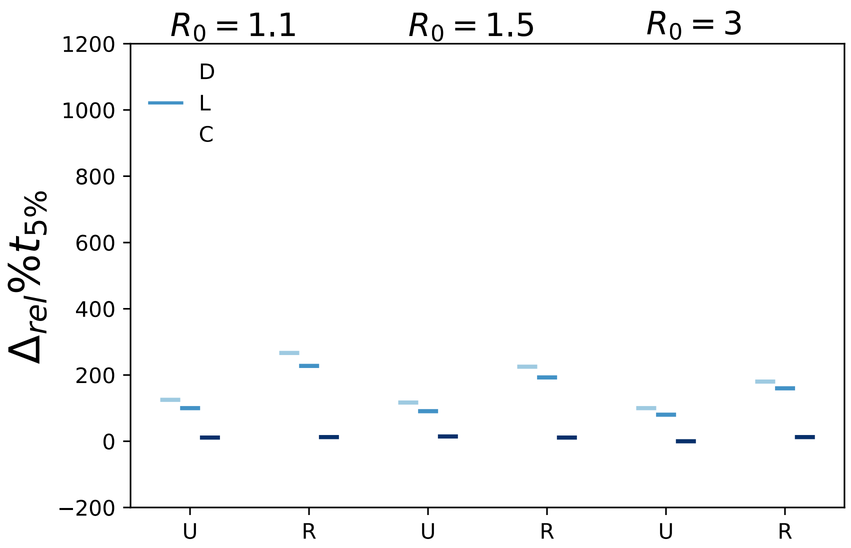


**Figure S4.** Relative variation of $t_{5\%}$. Box plots ranging from 5th to 95th percentile.

To better understand the differences between $D$,$L$,$C$ in the early-stage epidemics, here we present additional results on the time when the 5% of locations have been infected (**Figure S4**). As shows **Figure 6** the relative variation on $t_{5\%}$ between $L$ and $C$ do not exceed 200% and the median is equal to 0, while between $D$,$L$ and $D$,$C$ range from -100% to 1200%, with the median that is around 100%.

Complementary analysis

For the sake of completeness, in the SI are presented the complementary analysis relative to the main paper, showing all the epidemic parameters analyzed.

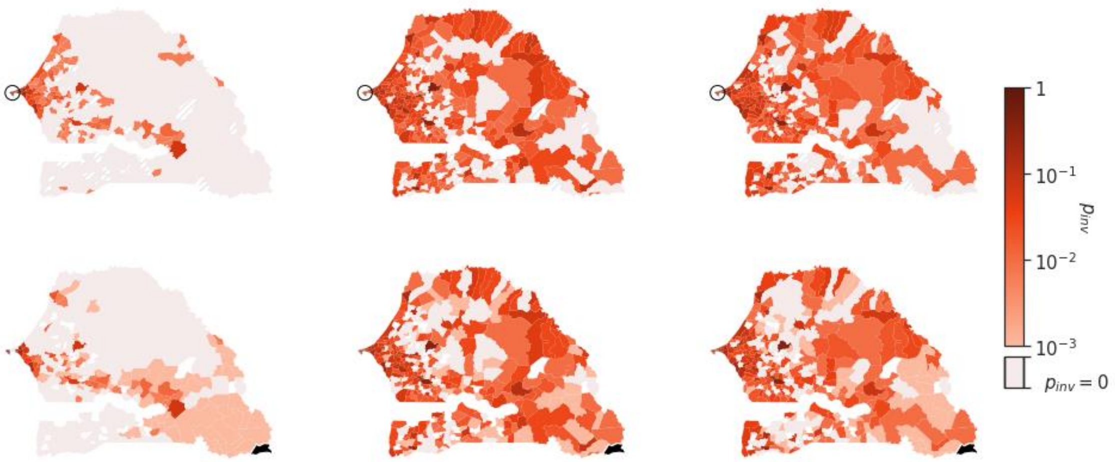


**Figure S5.** Map of the invasion probability at $t_{5\%}$ in any municipality with $R_0 = 1.1$. The municipality in black is the epidemic seed.

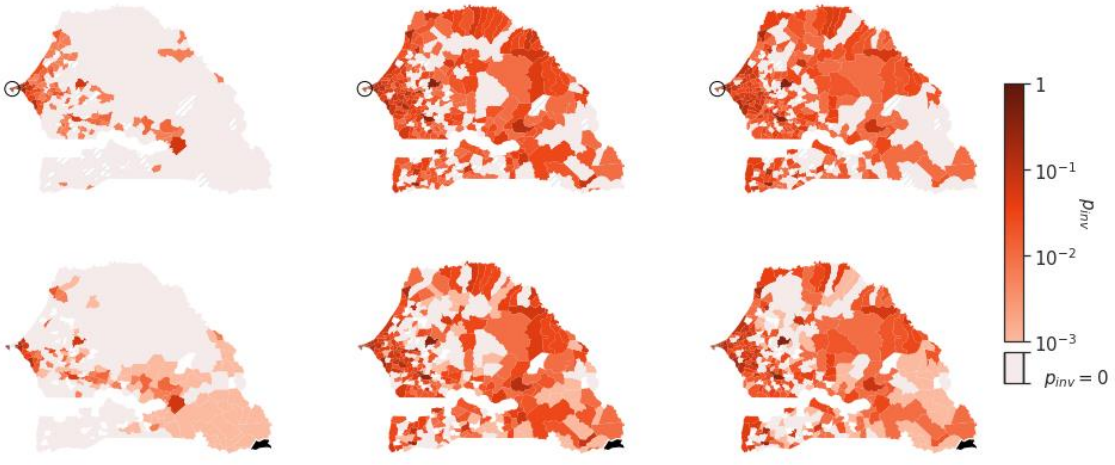


**Figure S5.** Map of the invasion probability at $t_{5\backslash\%}$ in any municipality with $R_0 = 1.5$. The municipality in black is the epidemic seed.

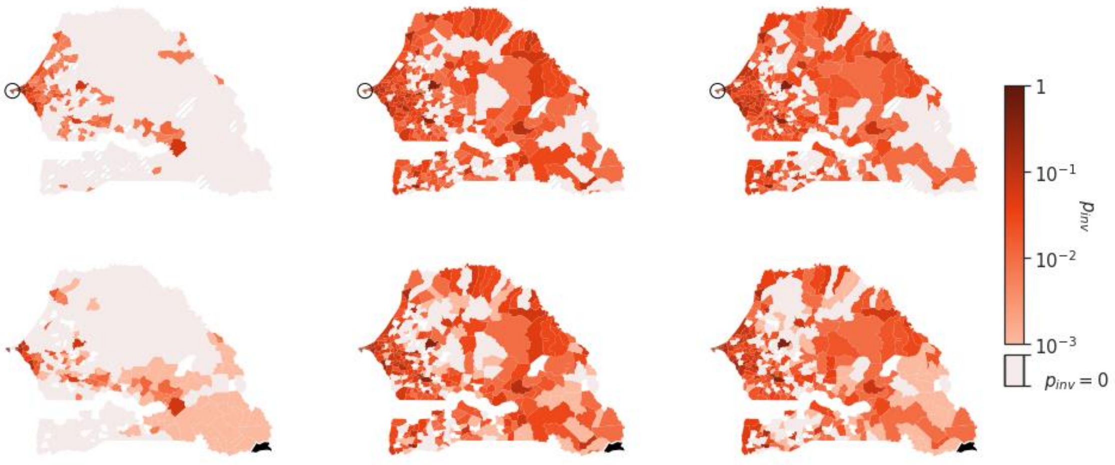


**Figure S5.** Map of the invasion probability at $t_{5\backslash\%}$ in any municipality with $R_0 = 3$. The municipality in black is the epidemic seed.

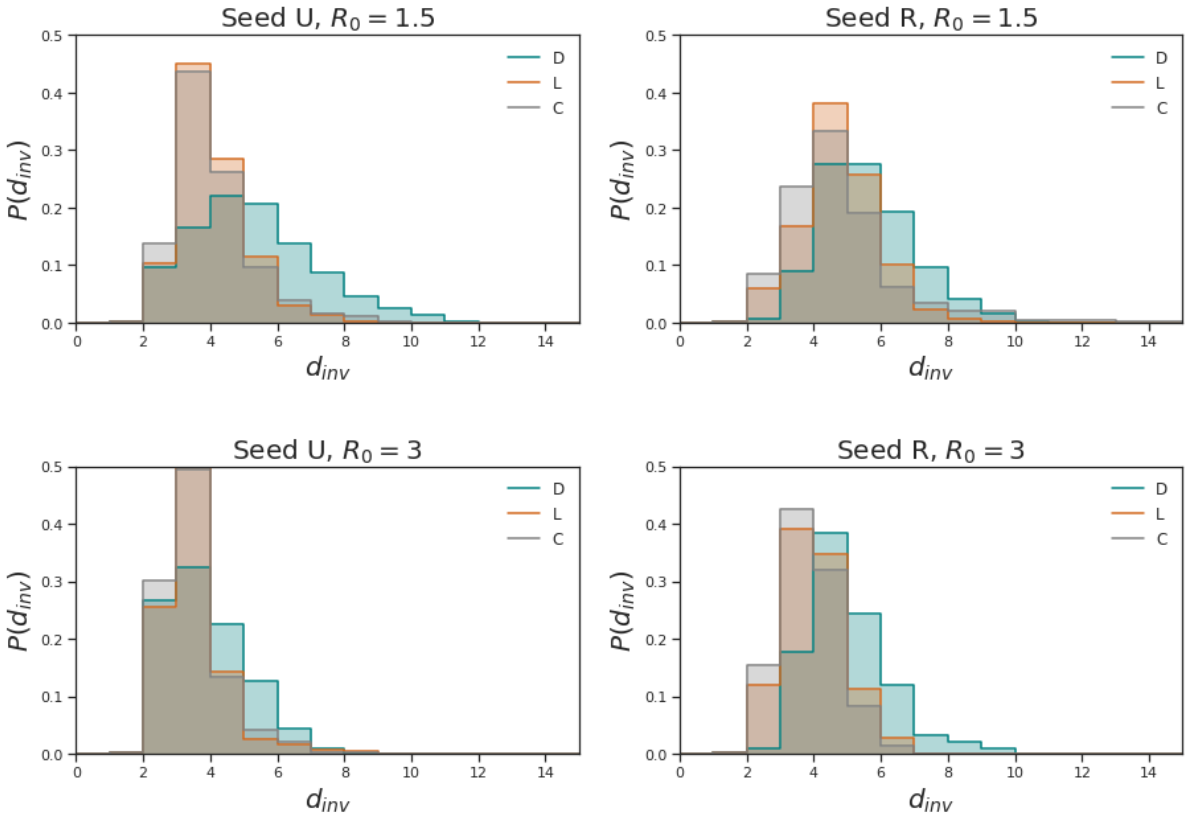


**Figure S6.** Invasion distance distributions in *D*, *L* and *C*

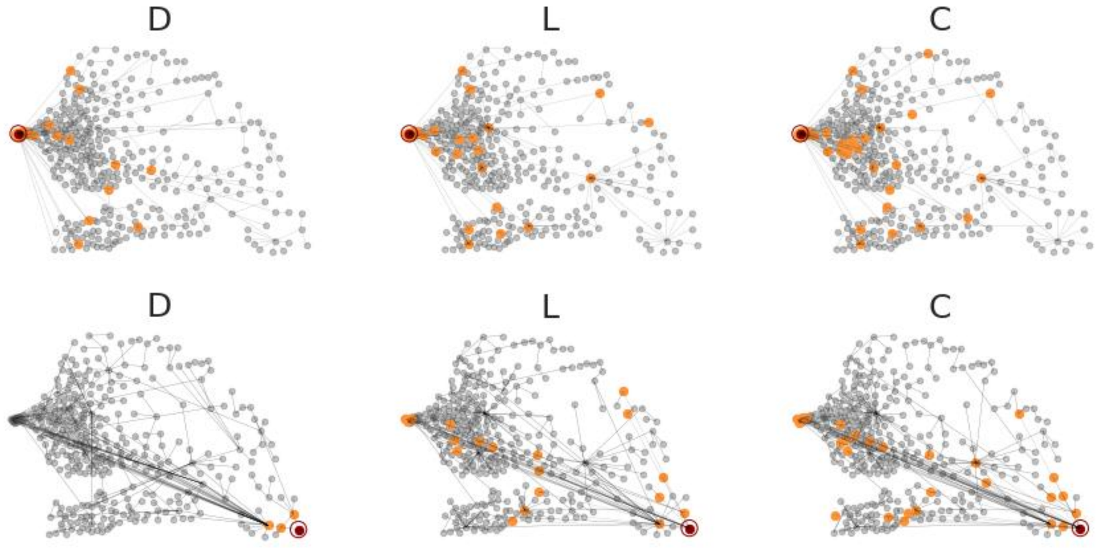


**Figure S7.** Invasion trees in *D, L*, *C* at $R_0 = 1.5$. The red dot is the epidemic seed, and the orange ones instead are the municipalities directly infected by the seed.

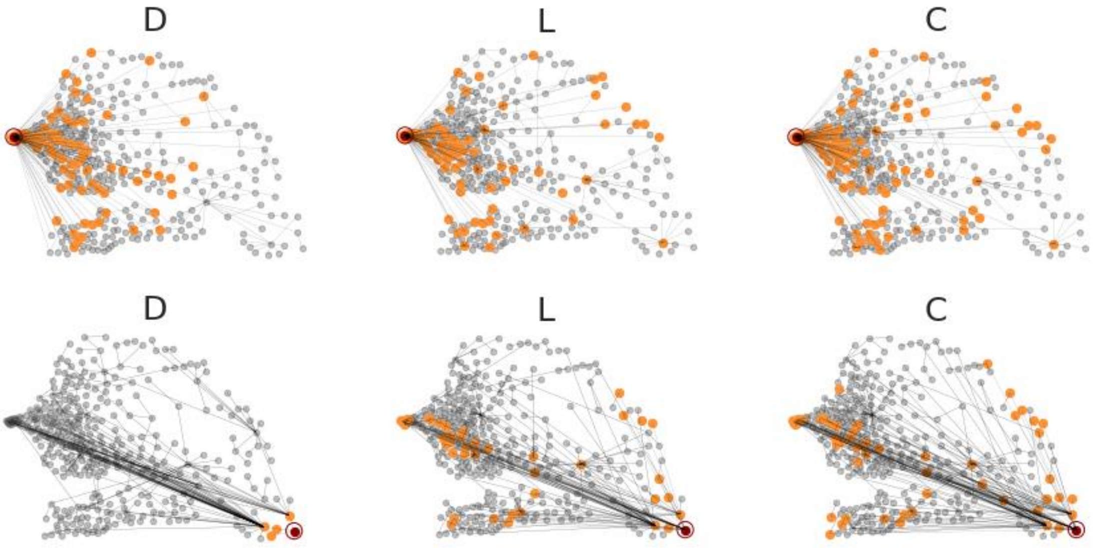


**Figure S8.** Invasion trees in $D$, $L$, $C$ at $R_0 = 3$. The red dot is the epidemic seed, and the orange ones instead are the municipalities directly infected by the seed.

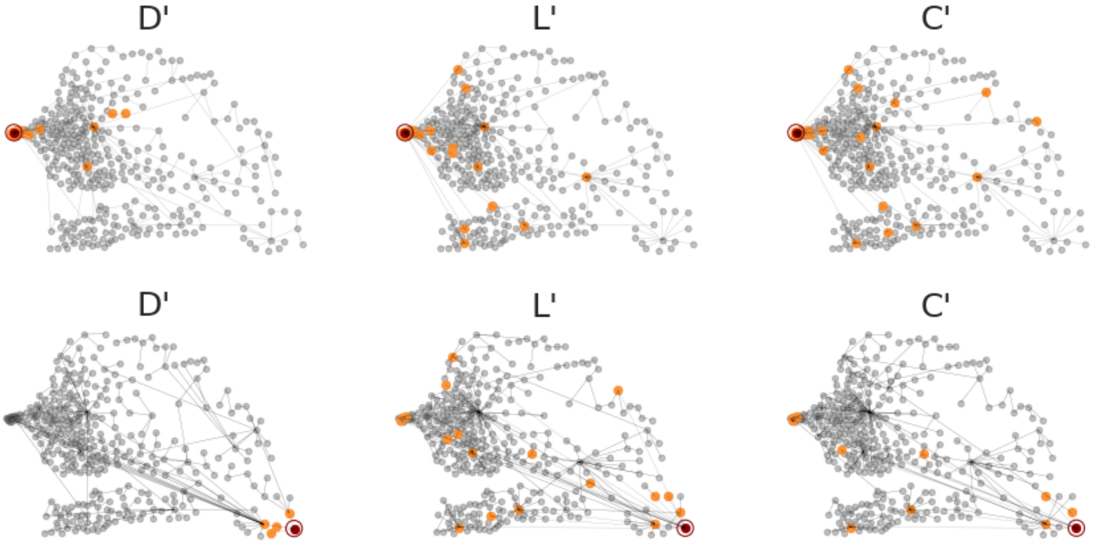


**Figure S9.** Invasion trees in $D'$, $L'$, $C'$ at $R_0 = 1.1$. The red dot is the epidemic seed, and the orange ones instead are the municipalities directly infected by the seed.

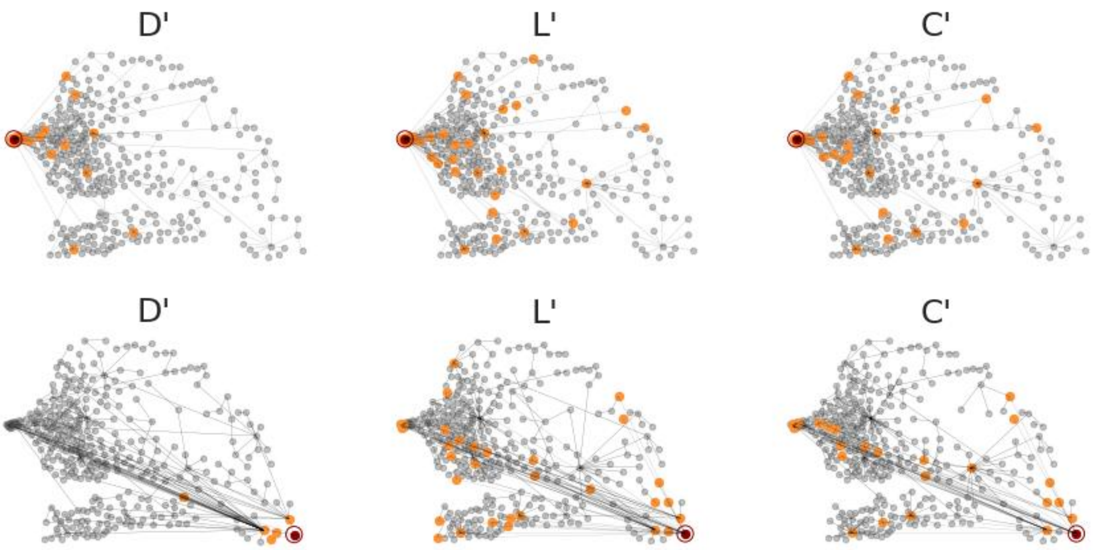


**Figure S10.** Invasion trees in $D'$, $L'$, $C'$ at $R_0 = 1.5$. The red dot is the epidemic seed, and the orange ones instead are the municipalities directly infected by the seed.

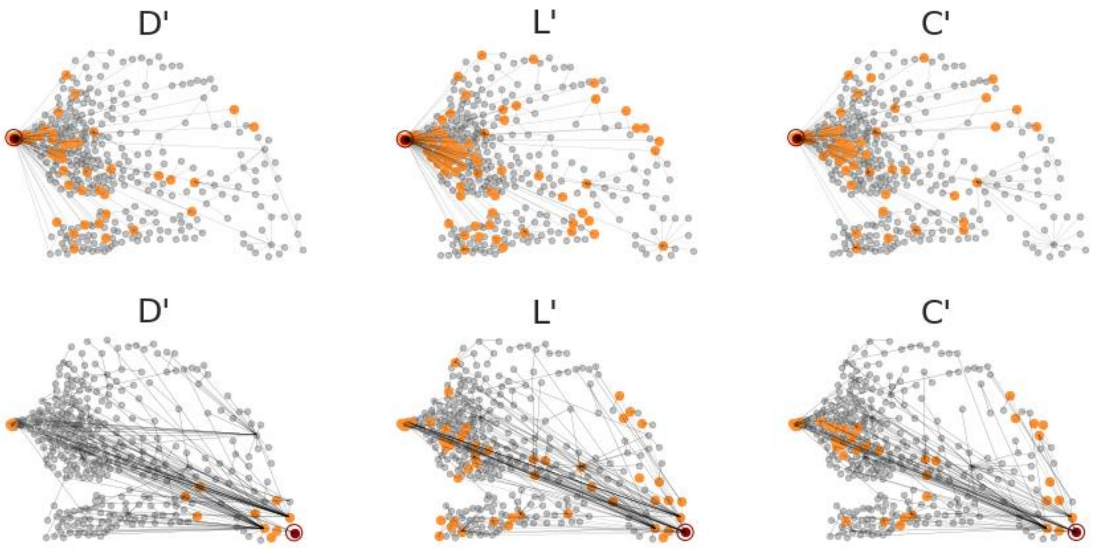


**Figure S11.** Invasion trees in $D'$, $L'$, $C'$ at $R_0 = 1.1$. The red dot is the epidemic seed, and the orange ones instead are the municipalities directly infected by the seed.

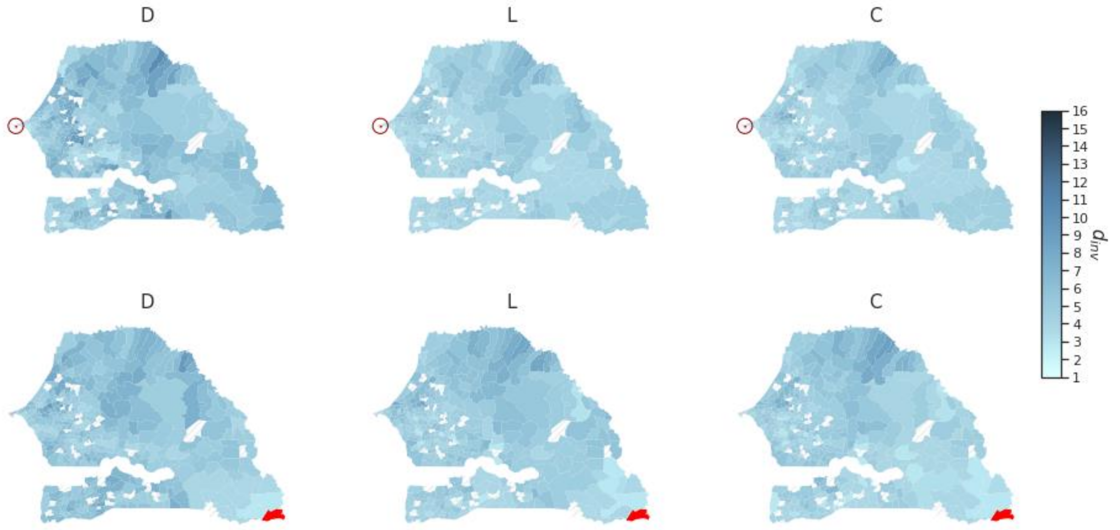


**Figure S12.** Visualization on map of the invasion distance at$R_0 = 1.5$ in $D$,$L$,$C$. The red municipality is the epidemic seed.

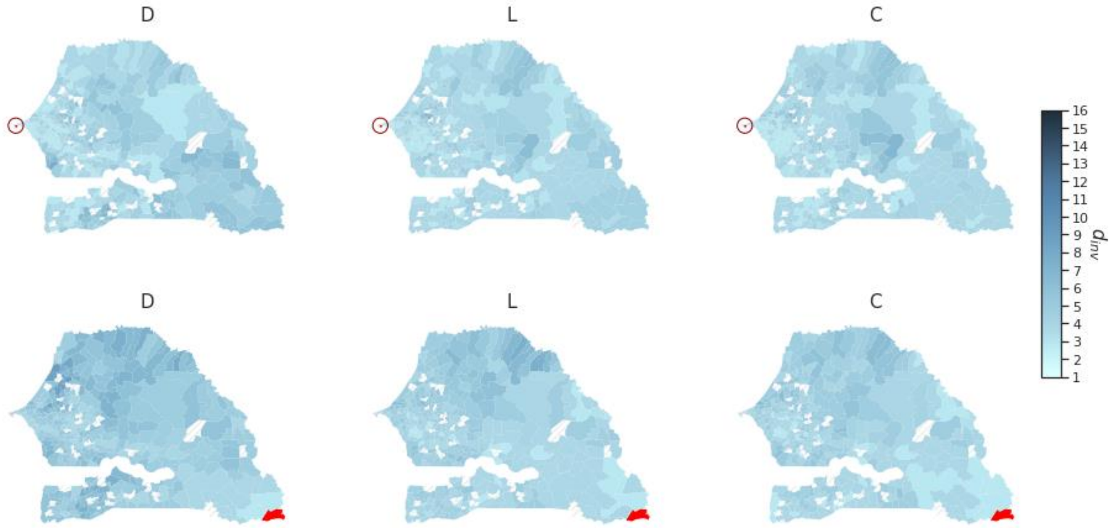


**Figure S13.** Visualization on map of the invasion distance at$R_0 = 1.5$ in $D$,$L$,$C$. The red municipality is the epidemic seed.

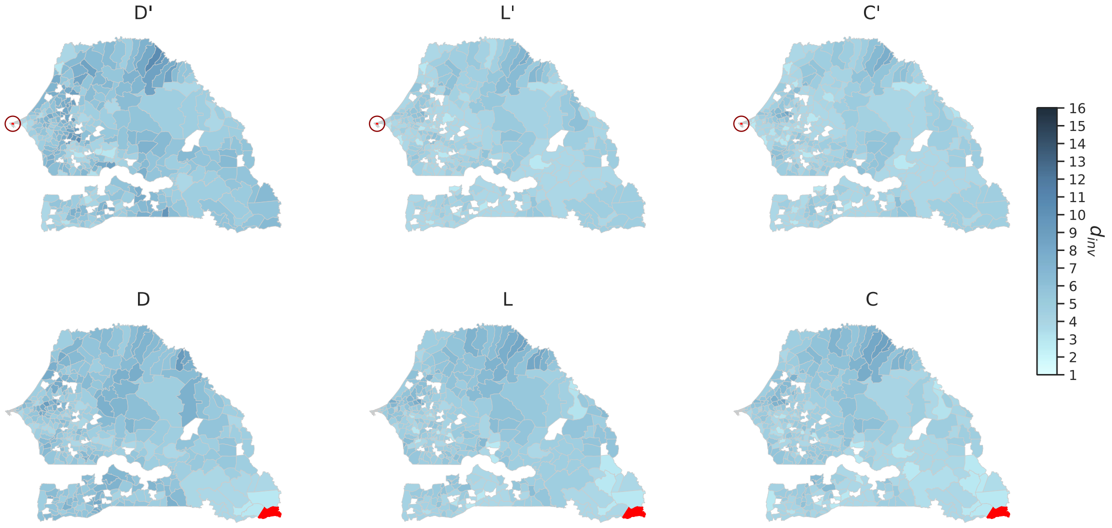


**Figure S14.** Visualization on map of the invasion distance at $R_0 = 1.5$ in $D'$, $L'$, $C'$. The red municipality is the epidemic seed.

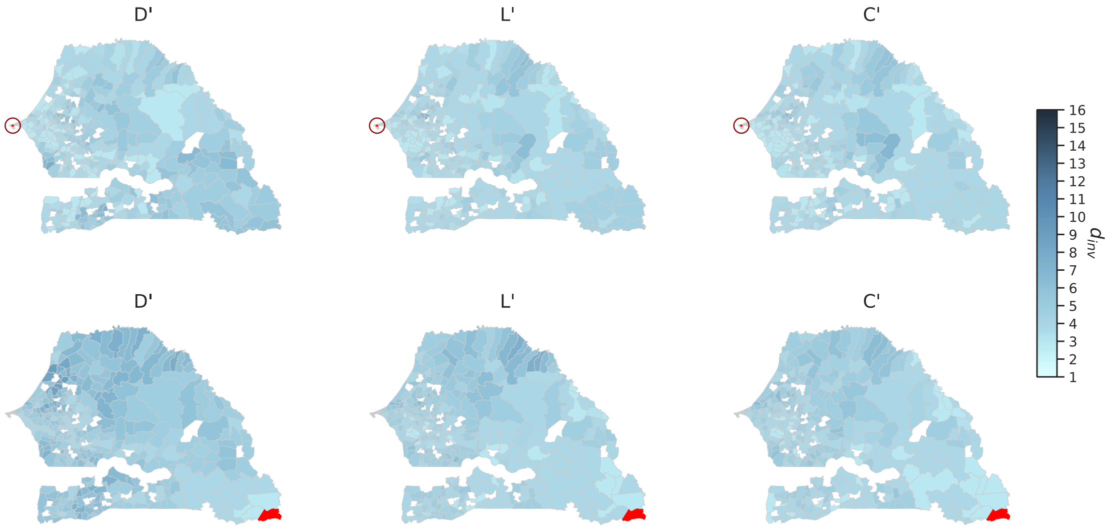


**Figure S15.** Visualization on map of the invasion distance at $R_0 = 3$ in $D'$, $L'$, $C'$. The red municipality is the epidemic seed.

<u>Correcting $D$</u>

We corrected the Displacement-matrix $D$ by defining $D_{norm}$. $D_{norm}$ takes into account the heterogeneity of the mobile phone users' activities (See Methods). Here we analyzed the differences between $D$ and $D_{norm}$ on comparing both the coupling matrices and the epidemic outcomes.

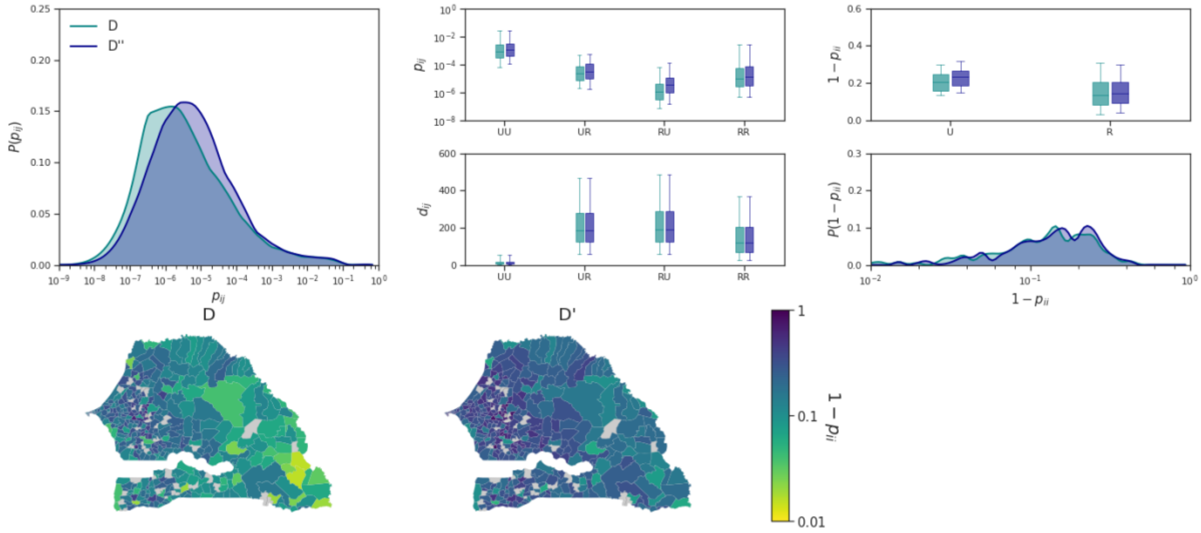


**Figure S16.** Differences between $D$ and $D_{norm}$. a) Distribution of the coupling probability in $D$ and $D_{norm}$. b),c),d) Coupling probability, Geographical distance and outgoing probability distribution in January of subset of links extracted by braking down municipalities in Urban (U) and Rural (R). e) Outgoing probability distribution in $D$ and $D_{norm}$. f) Visualization on map of the outgoing probability in $D$ and $D_{norm}$ per any municipality.
